# Exploring the Prevalence and Components of Metabolic Syndrome in Sub-Saharan African Type 2 Diabetes Mellitus Patients: A Systematic Review and Meta-Analysis

**DOI:** 10.1101/2024.02.15.24302899

**Authors:** Nelson Musilanga, Hussein Nasib, Given Jackson, Frank Shayo, Clarkson Nhanga, Saleh Girukwigomba, Ambokile Mwakibolwa, Samson Henry, Keneth Kijusya, Edgar Msonge

**Author notes:** Correspondence should be addressed to Nelson Musilanga.

## Abstract

**Background:** Type 2 diabetes mellitus and metabolic syndrome represent two closely intertwined public health challenges that have reached alarming epidemic proportions in low- and middle-income countries, particularly in sub-Saharan Africa. Therefore, the current study aimed to determine the weighted pooled prevalence of metabolic syndrome and its components among individuals with type 2 diabetes mellitus in sub-Saharan Africa as defined by the 2004 National Cholesterol Education Program-Adult Treatment Panel (NCEP-ATP III 2004) and/or the International Diabetes Federation (IDF) criteria.

**Methods:** A systematic search was conducted to retrieve studies published in the English language on the prevalence of metabolic syndrome among type 2 diabetic individuals in sub-Saharan Africa. Searches were carried out in PubMed, Embase, Scopus, Google Scholar, African Index Medicus and African Journal Online from their inception until July 31, 2023. A random-effects model was employed to estimate the weighted pooled prevalence of metabolic syndrome in sub-Saharan Africa. Evidence of between-study variance attributed to heterogeneity was assessed using Cochran’s Q statistic and the I2 statistic. The Joanna Briggs Institute quality appraisal criteria were used to evaluate the methodological quality of the included studies. The summary estimates were presented with forest plots and tables. Publication bias was checked with the funnel plot and Egger’s regression test.

**Results:** Overall, 1421 articles were identified and evaluated using the Preferred Reporting Items for Systematic Reviews and Meta-analyses (PRISMA) guidelines, and 30 studies that met the inclusion criteria were included in the final analysis. The weighted pooled prevalence of metabolic syndrome among individuals with type 2 diabetes mellitus in sub-Saharan Africa was 63.1% (95% CI: 57.9 – 68.1) when using the NCEP-ATP III 2004 criteria and 60.8% (95% CI: 50.7 – 70.0) when using the IDF criteria. Subgroup analysis, using NCEP-ATP III 2004 and IDF criteria, revealed higher weighted pooled prevalence among females: 73.5% (95% CI: 67.4-79.5), 71.6% (95% CI: 60.2-82.9), compared to males: 50.5% (95% CI: 43.8-57.2), 44.5% (95% CI: 34.2-54.8) respectively. Central obesity was the most prevalent component of metabolic syndrome, with a pooled prevalence of 55.9% and 61.6% using NCEP-ATP III 2004 and IDF criteria, respectively. There was no statistical evidence of publication bias in both the NCEP-ATP III 2004 and IDF pooled estimates.

**Conclusions:** The findings underscore the alarming prevalence of metabolic syndrome among individuals with type 2 diabetes mellitus in sub-Saharan Africa. Therefore, it is essential to promote lifestyle modifications, such as regular exercise and balanced diets, prioritize routine obesity screenings, and implement early interventions and robust public health measures to mitigate the risks associated with central obesity.

## Introduction

Metabolic Syndrome (MetS), characterized by a constellation of interconnected risk factors such as abdominal obesity, high blood pressure, high blood glucose, and abnormal lipid profiles, poses a significant risk to individuals worldwide [1,2]. When coexisting with type 2 diabetes mellitus (T2DM), this syndrome can exacerbate the progression of the disease and increase the risk of cardiovascular diseases [3,4], which are the leading cause of mortality worldwide [5,6]. Sub-Saharan Africa (SSA), home to over one billion people, is not immune to these global health trends [7]. Owing to the increase in urbanization, excessive alcohol consumption, unhealthy eating habits, smoking, sedentary lifestyles, and overweight [8,9], SSA like many other regions, is currently witnessing a rapid epidemiological shift characterized by an increasing predominance of non-communicable diseases (NCDs) [10], contributing to a growing prevalence of both T2DM and MetS in the region.

T2DM is the most common chronic metabolic-endocrine disorder affecting adults. It results from a complex interaction between heredity along with other risk factors such as insulin resistance, obesity, physical inactivity, an unhealthy diets, smoking, and excessive alcohol consumption [11]. It’s multi-systemic nature suggests that complications and comorbidities have the potential to impact various organ systems [12], particularly in the setting of poor blood glucose control. The burden of T2DM in sub-Saharan Africa has grown into a substantial public health challenge. According to the International Diabetes Federation (IDF) report, the greatest relative increase in the prevalence of diabetes between 2021 and 2045 will occur in low-income countries (11.9%) and middle-income countries (21.1%), which largely includes SSA countries [13].

Globally, the prevalence of MetS is escalating at an alarming rate, and it is highly prevalent in patients with T2DM [14,15]. It was estimated that 20% to 25% of the adult general population and 70% to 80% of T2DM patients had MetS worldwide [16]. Individuals with MetS are more likely to have a higher risk of heart attacks and cardiovascular diseases (CVD) compared to those without MetS [4]. Furthermore, it is documented that the risk of CVD development is greater among individuals who have a combination of T2DM and MetS compared to those who have either condition alone [17].

While the burden of communicable diseases has traditionally been the major focus of public health initiatives in SSA, the rise of non-communicable diseases like T2DM and MetS is now posing a significant threat to the region’s health and socio-economic development. Unlike prior studies [18,19] that explored MetS in broader African populations or specific country, the current study aimed to systematically review the available evidence and provide an estimate of the pooled prevalence of MetS among SSA individuals with T2DM. By spotlighting MetS within the context of T2DM in SSA, offers a more targeted understanding of MetS within a unique subset of the African population, providing valuable information for healthcare practitioners and researchers focusing on this demographic.

## Methods

### Design and Registration

We conducted a systematic review and meta-analysis of observational studies, all of which were cross-sectional study designs done across SSA. This systematic review and meta-analysis was reported according to the Preferred Reporting Items for Systematic Reviews and Meta-Analyses (PRISMA) statement guideline [20]. The study protocol was registered in the PROSPERO, an international prospective register of systematic reviews protocols on health related topics https://www.crd.york.ac.uk/PROSPERO/display_record.php?RecordID=455576CRD42023455576 [21].

### Outcome of Interest

The primary outcome of interest for this study was the pooled prevalence of MetS among T2DM patients, as defined by the widely recognized and extensively used criteria’s i.e 2004 National Cholesterol Education Program-Adult Treatment Panel (NCEP-ATP III 2004) [1] and/or the IDF criteria [2]. Using NCEP-ATP III 2004 any three of the five metabolic syndrome components while using IDF criteria central Obesity, plus two of the four MetS components (table 1). The secondary aim was to describe the prevalence of individual components of MetS among T2DM patients, according to the specific MetS definition criteria among T2DM individuals in SSA.

**Table 1:** Diagnostic criteria of metabolic syndrome according to NCEP-ATP III 2004 and IDF Criteria.

| Criteria | NCEP-ATP III 2004 | IDF |
| --- | --- | --- |
| Central Obesity | Waist circumference $\geq 102$cm in male and $\geq 88$cm in female. | Waist circumference $\geq 94$cm in male and $\geq 80$cm in female. |
| Hypertriglyceridemia | TG $\geq 150$ mg/dl or triglycerides treatment | TG $\geq 150$ mg/dl or triglycerides treatment |
| Reduced HDL-Cholesterol | $<40$mg/dl in males and $<50$mg/dl in females or HDL-c treatment | $<40$mg/dl in males and $<50$mg/dl in females or HDL-c treatment |
| Hyperglycemia | FBG $\geq 100$mg/dL or on treatment | FBG $\geq 100$mg/dL or on treatment |
| Hypertension | Systolic/Diastolic BP $\geq 130/85$mmHg or hypertension treatment | Systolic/Diastolic BP $\geq 130/85$mmHg or hypertension treatment |

BP: Blood pressure; FBG: Fasting blood glucose; HDL-c: High Density Lipoprotein cholesterol; IDF: International Diabetes Federation; NCEP-ATP III: National Cholesterol Education Program–Adult Treatment Panel III; TG: Triglyceride.

### Data Source and Search Strategy

We conducted a comprehensive systematic literature search to identify studies reporting the prevalence of MetS among T2DM patients in the sub-Saharan African population. The search utilized a combination of Medical Subject Headings (MeSH) and free text words across various electronic databases and search engine, including MEDLINE-PubMed, EMBASE, Scopus, African Index Medicus, African Journal Online and Google Scholar. Inclusion criteria were limited to English-language studies published from the inception of databases until July 31, 2023. Additionally, a snowball search was performed on the reference lists of all relevant included studies. The search strategy focused on three key elements: metabolic syndrome, type 2 diabetes mellitus and sub-Saharan Africa. These searches were independently performed by two authors; N.M and H.N. The detailed search strategy used for the databases is presented in the supplementary material S1. To manage references and remove duplicates, we used Rayyan, an online web application.

### Inclusion and exclusion criteria

The inclusion criteria were as follows: All observational studies reporting the prevalence of MetS and its subcomponents among T2DM individuals in sub-Saharan African populations, studies reporting metabolic syndrome using IDF criteria and/or NCEP-ATP III 2004, and publications with full text in English. The full text of studies meeting these criteria was retrieved and screened for eligibility. Whereas, non-original research articles, such as review articles, editorials, case reports, letters, or commentaries, studies describing MetS in populations other than sub-Saharan Africa, T2DM, and those with unclear or unspecified methods of diagnosing metabolic syndrome were excluded.

### Study Selection and Quality Assessment

Two authors (N.M. and H.N.) independently conducted the literature search and screened the titles, abstracts and keywords of all the studies retrieved from online database searches for possible inclusion in the review. Furthermore, the relevant articles were obtained in full text, and after a thorough reading of the full-text articles, the included studies were identified based on the assessment of inclusion and exclusion criteria. Any discrepancies during the entire selection process between the two authors were resolved either through consensus or consultation with third author (G.J). The search, screening, and study identification process are summarized in Fig. 1. The methodological quality and risk of bias of the included studies was assessed using eight aspects of the Joanna Brigg’s Institute (JBI) quality checklist for analytical cross-sectional studies [22,23]. Two authors (N.M. and H.N.) independently used the tool to evaluate the inclusion criteria, measurement of exposure and outcome variables, confounding adjustment, and appropriateness of statistical analysis. Studies that scored 50% or higher on the quality assessment were considered to be of good quality. Full details regarding the appraisal checklist are provided in table 2.

**Figure 1:**
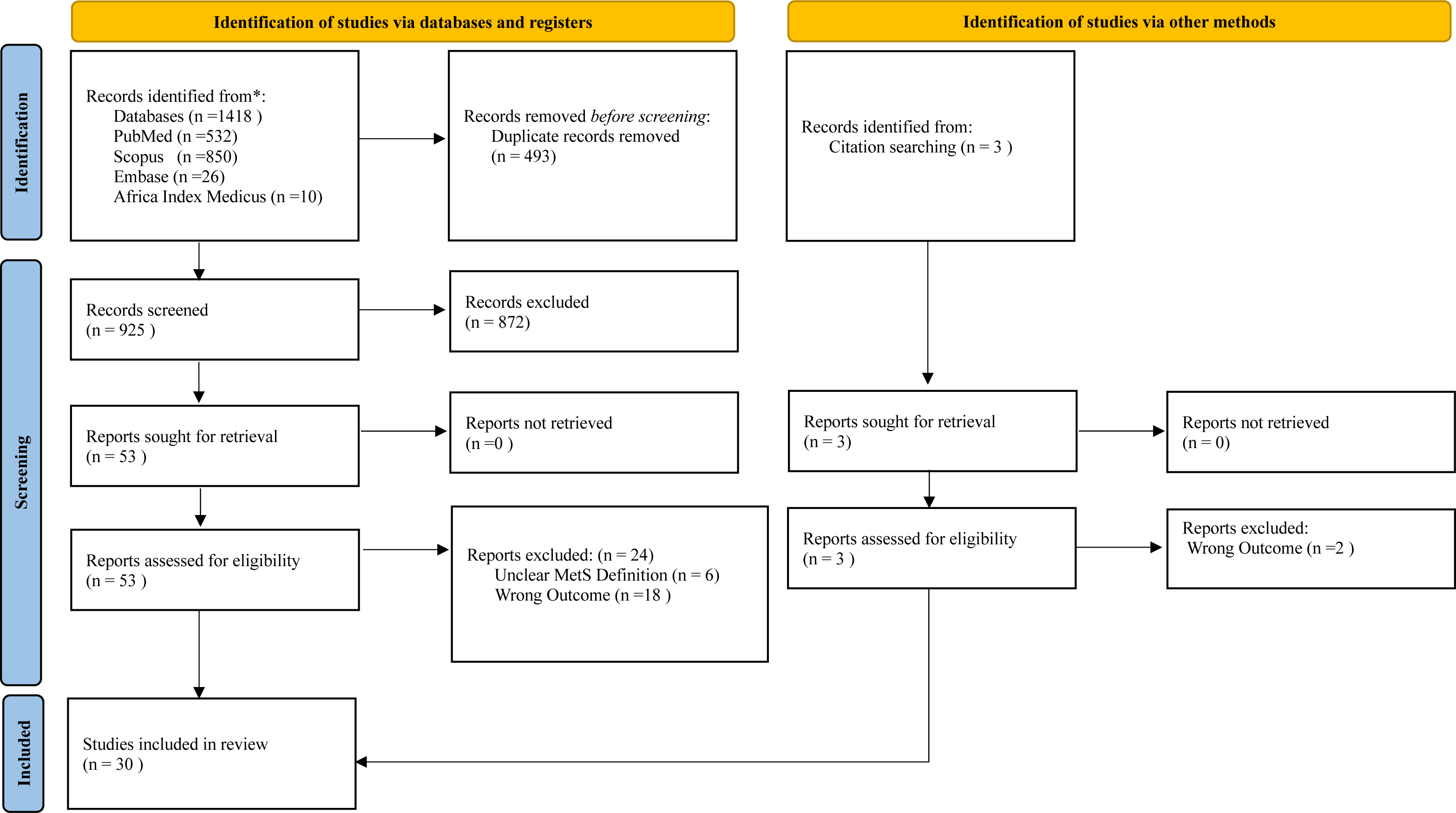
Preferred reporting items for systematic reviews and meta-analyses (PRISMA) flow chart.

**Table 2:** Methodological quality assessment of included studies using Joanna Brigg’s Institute quality appraisal (JBI)

| Author[Year] | Were the criteria for inclusion in the sample clearly defined? | Were the study participants and setting described in detail? | Was the exposure measured in a valid and reliable way? | Were objective, standard criteria used for measurement of the condition? | Was appropriate statistical analysis used? | Were the outcomes measured in a valid and reliable way? | Were confounding factors identified? | Were strategies to deal with confounding factors stated? | Overall appraisal |
| --- | --- | --- | --- | --- | --- | --- | --- | --- | --- |
| Kalk et al. [24] 2008 | Yes | Yes | Yes | Yes | Yes | Yes | Yes | Yes | Good |
| Titty et al. [25] 2008 | Yes | Yes | Yes | Yes | Yes | Yes | Unclear | Unclear | Good |
| Titty et al. [26] 2009 | Yes | Yes | Yes | Yes | Yes | Yes | Unclear | Unclear | Good |
| Puepet et al. [27] 2009 | Yes | Yes | Yes | Yes | Yes | Yes | Unclear | Unclear | Good |
| Unadike et al. [28] 2009 | Yes | Yes | Yes | Yes | Yes | Yes | Unclear | Unclear | Good |
| Chanda et al. [29] 2010 | Yes | Yes | Yes | Yes | Yes | Yes | Unclear | Unclear | Good |
| Titty et al. [30] 2010 | Yes | Yes | Yes | Yes | Yes | Yes | Unclear | Unclear | Good |
| Ogbera et al. [31] 2011 | Yes | Yes | Yes | Yes | Yes | Yes | Yes | Yes | Good |
| Kangne et al. [32] 2012 | Yes | Yes | Yes | Yes | Yes | Yes | Yes | Yes | Good |
| Osuji et al. [33] 2012 | Yes | Yes | Yes | Yes | Yes | Yes | Unclear | Unclear | Good |
| Mogre et al. [34] 2014 | Yes | Yes | Yes | Yes | Yes | Yes | Yes | Yes | Good |
| Nsiah et al. [35] 2015 | Yes | Yes | Yes | Yes | Yes | Yes | Yes | Yes | Good |
| Ejiofor et al. [36] 2015 | Yes | Yes | Yes | Yes | Yes | Yes | Unclear | Unclear | Good |
| Onyenekwu et al. [37] 2017 | Yes | Yes | Yes | Yes | Yes | Yes | Unclear | Unclear | Good |
| Abban et al. [38] 2017 | Yes | Yes | Yes | Yes | Yes | Yes | Yes | Yes | Good |
| Amidu et al. [39] 2017 | Yes | Yes | Yes | Yes | Yes | Yes | Unclear | Unclear | Good |
| Osei-Yeboah et al. [40] 2017 | Yes | Yes | Yes | Yes | Yes | Yes | Yes | Yes | Good |
| Woyesa et al. [41] 2017 | Yes | Yes | Yes | Yes | Yes | Yes | Yes | Yes | Good |
| Tadewos et al. [42] 2017 | Yes | Yes | Yes | Yes | Yes | Yes | Yes | Yes | Good |
| Biadgo et al. [43] 2018 | Yes | Yes | Yes | Yes | Yes | Yes | Yes | Yes | Good |
| Birarra et al. [44] 2018 | Yes | Yes | Yes | Yes | Yes | Yes | Yes | Yes | Good |
| Obirikorang et al. [45] 2018 | Yes | Yes | Yes | Yes | Yes | Yes | Yes | Yes | Good |
| Agyemang-Yeboah et al. [46] 2019 | Yes | Yes | Yes | Yes | Yes | Yes | Yes | Yes | Good |
| Gebremeskel et al. [47] 2019 | Yes | Yes | Yes | Yes | Yes | Yes | Yes | Yes | Good |
| Wube et al. [48] 2019 | Yes | Yes | Yes | Yes | Yes | Yes | Yes | Yes | Good |
| Zerga et al. [49] 2020 | Yes | Yes | Yes | Yes | Yes | Yes | Yes | Yes | Good |
| Anto et al. [50] 2022 | Yes | Yes | Yes | Yes | Yes | Yes | Yes | Yes | Good |
| Gebreyesus et al. [51] 2022 | Yes | Yes | Yes | Yes | Yes | Yes | Yes | Yes | Good |
| Gemeda et al. [52] 2022 | Yes | Yes | Yes | Yes | Yes | Yes | Yes | Yes | Good |
| Charkos et al. [53] 2023 | Yes | Yes | Yes | Yes | Yes | Yes | Yes | Yes | Good |

### Data Extraction

Extraction of relevant data from the included studies was independently performed by two authors (N.M and H.N). Information regarding authors; year of publication; geographical location; year(s) of survey; study design; sample size; gender; mean age; sampling techniques; diagnostic criteria for defining metabolic syndrome and relevant clinic outcomes of interest were collected using a standardized data extraction form. Extracted data were then checked for its accuracy and consistency by a third author (G.J).

### Statistical Analysis

The extracted data were exported to computer software RStudio version 2023.06.1+524 for data synthesis, analysis, and generation of forest and funnel plots. Evidence of between study variance due to heterogeneity was assessed using Cochran’s Q statistic and the *I^2^* statistic [54,55]. Furthermore, in order to explore potential sources of heterogeneity across the included studies, subgroup and sensitivity analyses were performed to comprehensively assess the overall effect size within the included studies. A random-effects model with inverse variance was used to obtain an overall summary estimate of the prevalence across studies. Point estimation with a confidence interval of 95% was used. The presence of publication bias was examined through the utilization of funnel plots, further enhanced by formal statistical assessment using Egger’s test [56].

## Results

### Study Selection

As shown in figure 1, a preliminary search of online databases using a combination of MeSH and free text words retrieved a total of 1418 potential articles, and an additional 3 articles were found through manual citation searching. After removing duplicates, 928 articles remained, which were then screened based on their titles and abstracts, resulting in the elimination of a further 872 articles that were irrelevant to the research question. Among the 56 articles that underwent full-text review, ultimately 30 articles met the inclusion criteria and were included in this review.

### Characteristics of Included Studies

A characteristic summary of thirty articles included in this study involving 8879 individuals is illustrated in table 3. All were of cross-sectional study design conducted in six sub-Saharan African countries namely Cameroon, Ethiopia, Ghana, Nigeria, Zambia and South Africa as demonstrated in figure 2. In these studies, the prevalence of MetS was estimated based on the IDF and/or NCEP-ATP III 2004 criteria. Among the articles, eleven studies reported the prevalence of MetS based on both NCEP-ATP III 2004 and IDF criteria [32,38,53,39,40,43–45,48,49,51], fourteen studies reported based solely on NCEP-ATP III 2004 criteria [25,26,42,46,50,52,28–31,33,35,36,41] and five studies reported based on IDF criteria alone [24,27,34,37,47]. Additionally, nine studies reported the prevalence of MetS subcomponents based on NCEP-ATP III 2004 criteria [25,26,31,35,40–44] and six studies based on IDF criteria [24,27,40,43,44,47].

**Figure 2:**
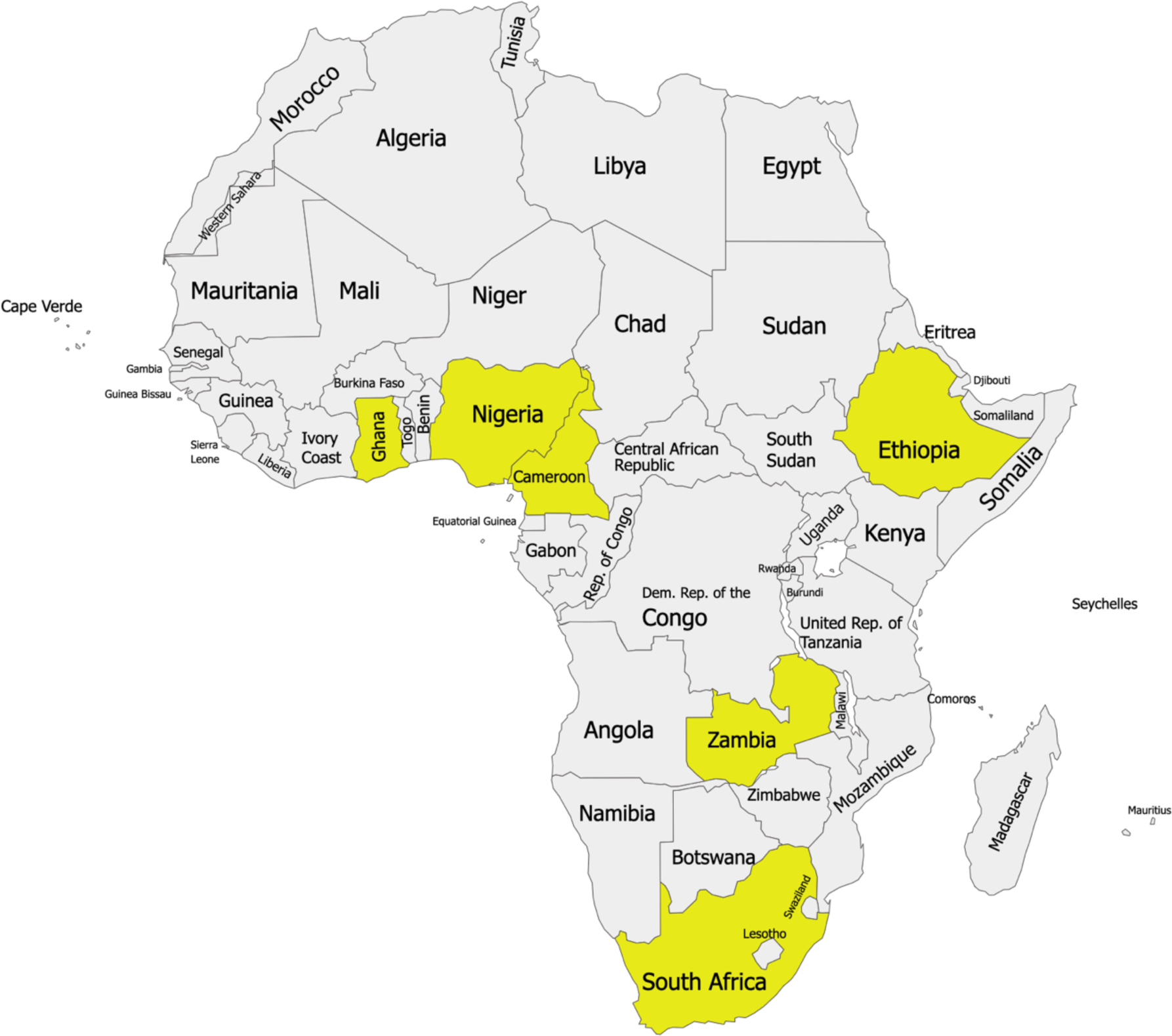
A map of Africa showing the locations of the included studies (created with paintmaps.com)

**Table 3:**
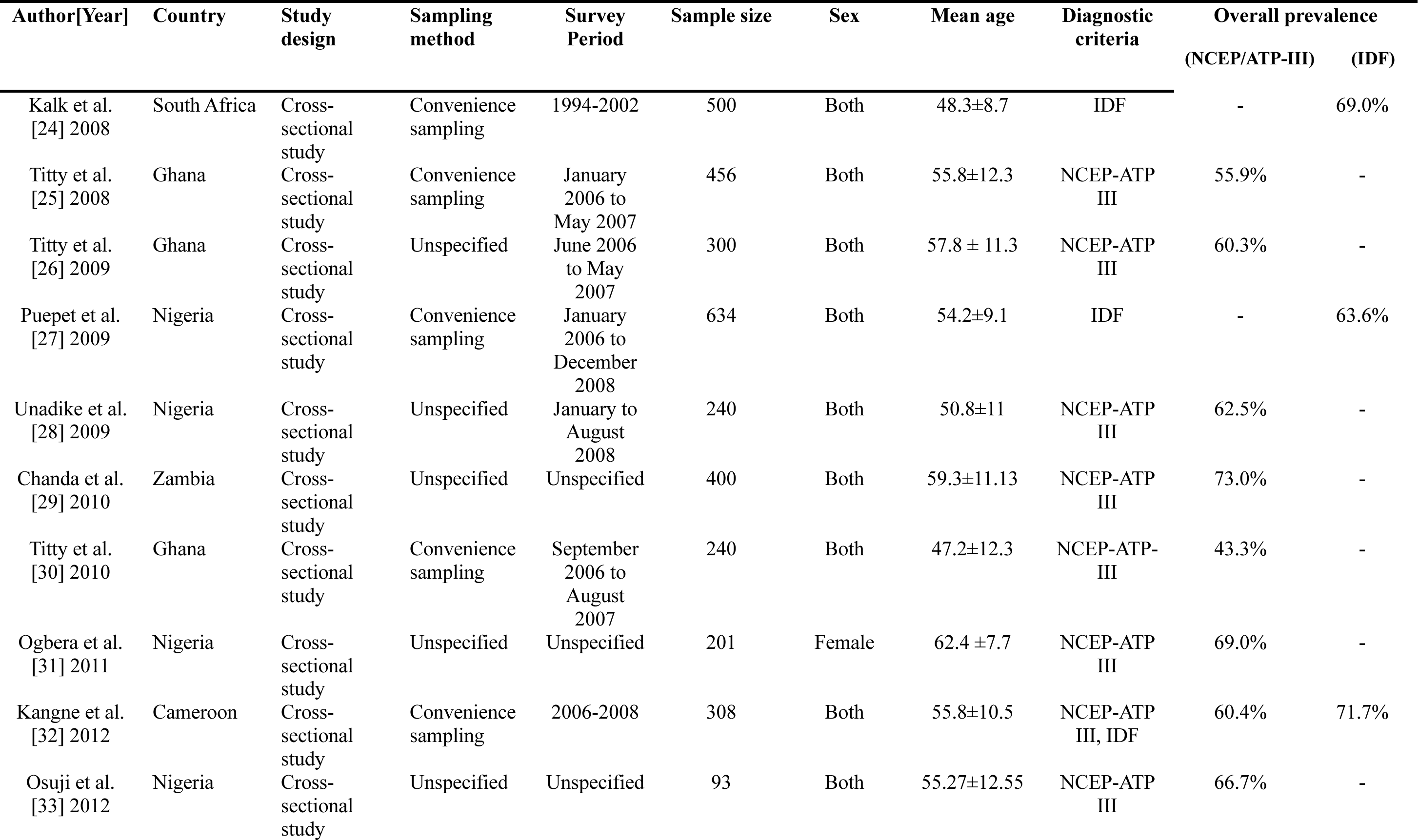
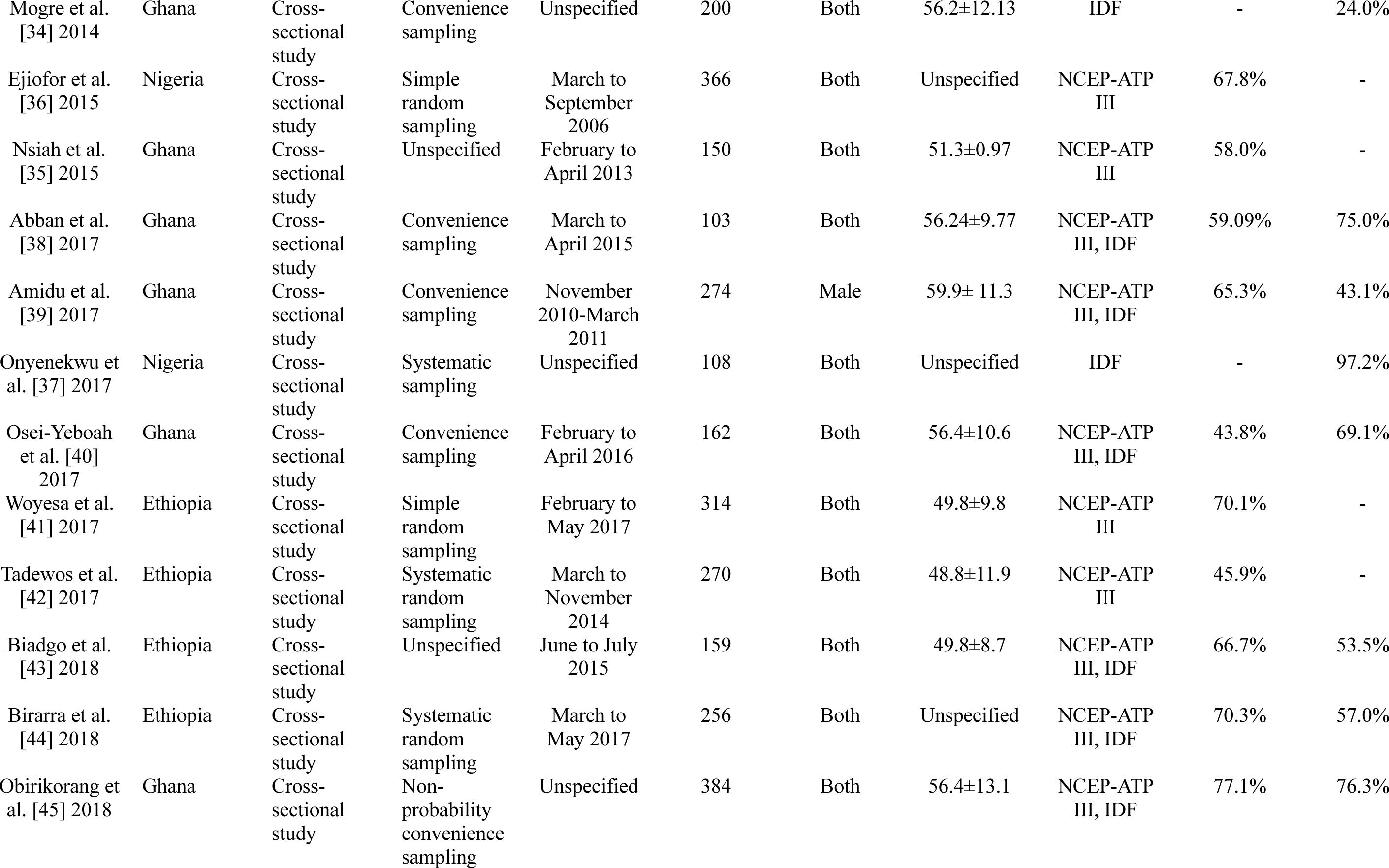
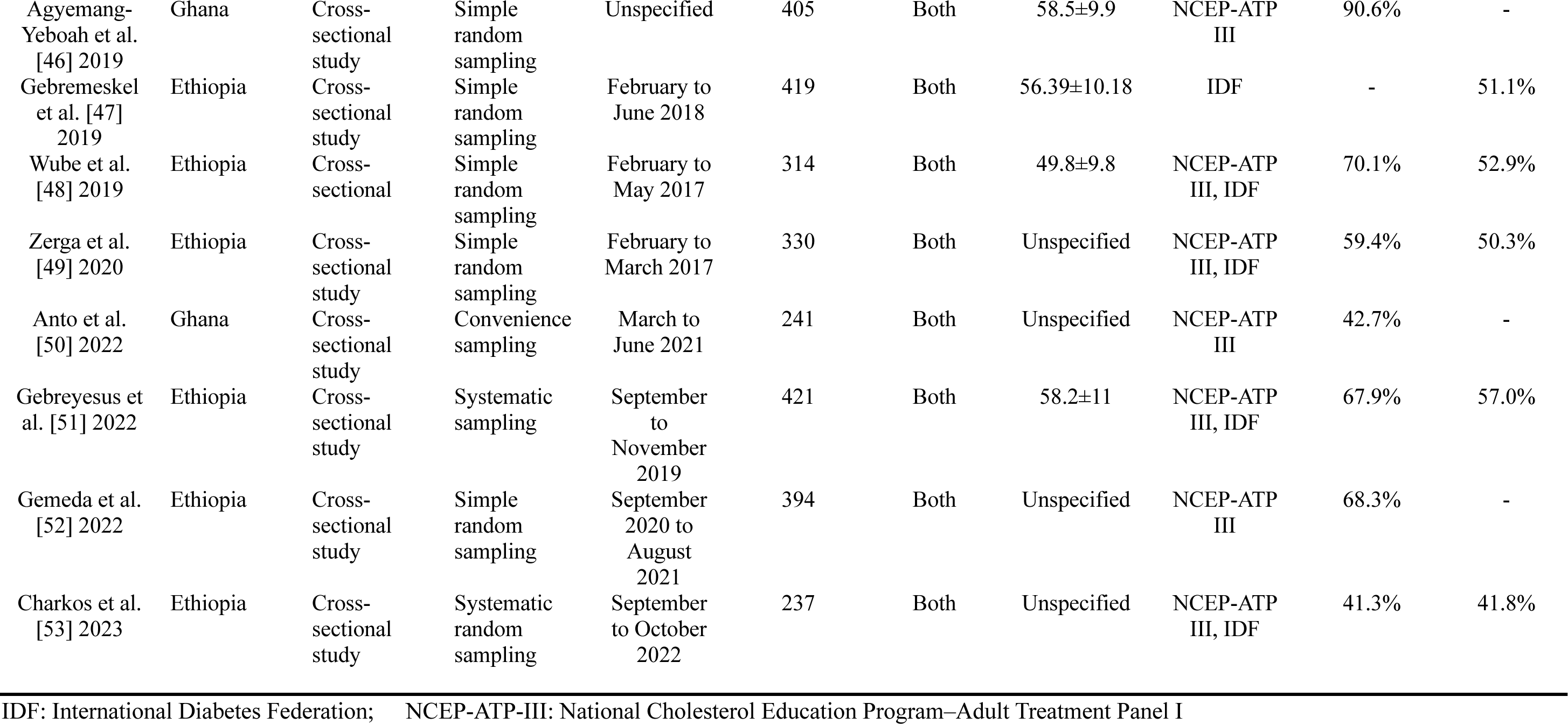
Characteristics of the included studies that evaluated the prevalence of MetS among T2DM in sub-Saharan population.

### Burden of Metabolic syndrome Using NCEP-ATP III 2004 and IDF Criteria

The weighted pooled prevalence of MetS among T2DM individuals in sub-Saharan Africa using NCEP-ATP III 2004 criteria is 63.1% (95% CI: 57.9 – 68.1), with significant heterogeneity *I^2^*=94% and Cochran Q-statistic p < 0.01 as graphically depicted in figure 3. While using IDF criteria yielded a pooled prevalence of 60.8% (95% CI: 50.7 – 70.0), with an *I^2^* of 95% and Cochran Q-statistic p < 0.01 as shown in figure 4. The random-effects model was assumed due to the considerable heterogeneity observed across the included studies in the meta-analysis.

**Figure 3:**
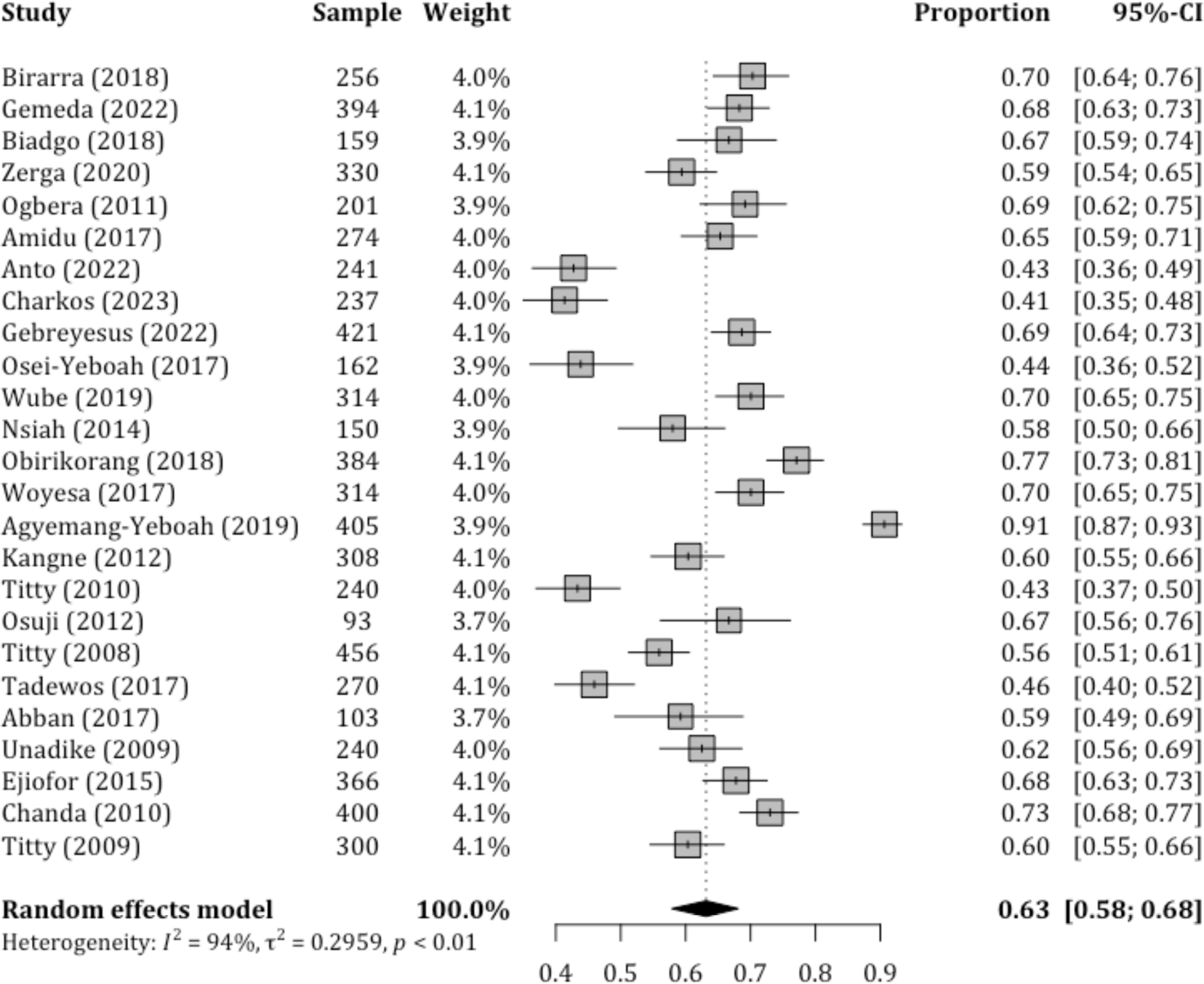
Forest plot illustrating the pooled prevalence of MetS with corresponding 95% CIs in sub-Saharan Africa based on NCEP-ATP III 2004 criteria.

**Figure 4:**
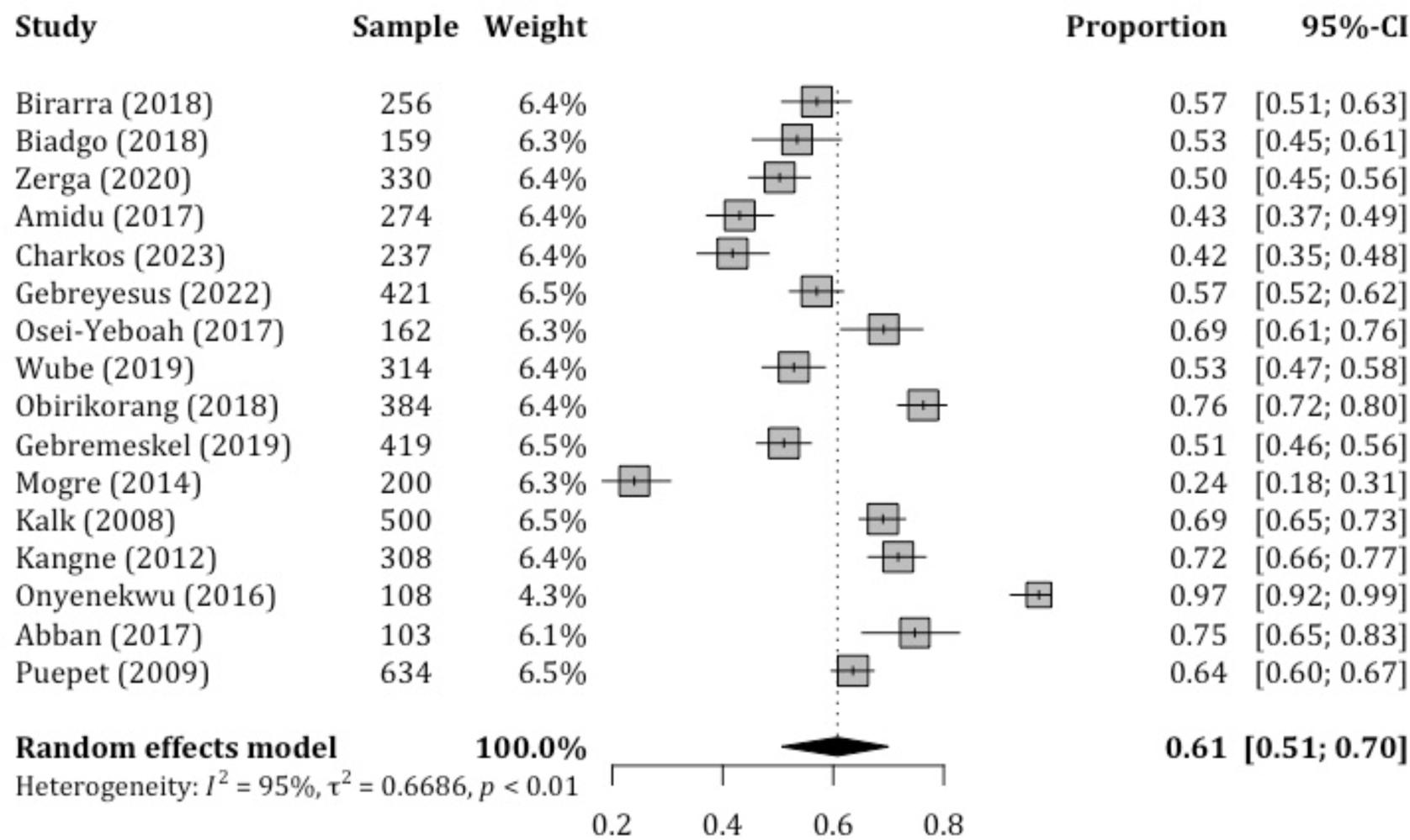
Forest plot illustrating the pooled prevalence of MetS with corresponding 95% CIs in sub-Saharan Africa based on IDF criteria.

### Prevalence of the Metabolic Syndrome Components

In the current systematic review, the prevalence of the individual components of MetS other than hyperglycemia among the sub-Saharan Africa T2DM population was reported in ten studies based on NCEP-ATP III 2004 criteria, and six studies reported based on IDF criteria. The overall pooled prevalence of metabolic syndrome component by NCEP-ATP III 2004 criteria was as follows: central obesity 55.9% [95% CI: 47.6, 64.2], low HDL-c 43.3% [95% CI: 33.5, 53.2], hypertriglyceridemia 48.0% [95% CI: 35.2, 60.7] and hypertension 54.8% [95% CI: 43.2, 66.4]. These values are summarized in table 4.

**Table 4:** Pooled prevalence of metabolic syndrome component based on NCEP-ATP III 2004.

| Prevalence of metabolic syndrome component |  |  |  |  |  |
| --- | --- | --- | --- | --- | --- |
| Author [Year] | Sample | Central Obesity | Low-HDL-c | High-TG | Hypertension |
| Titty et al. [25] 2008 | 456 | 43.6 | 47.4 | 37.5 | 46.9 |
| Titty et al. [26] 2009 | 300 | 69.6 | 58.5 | 56.4 | 69.6 |
| Unadike et al. [28] 2009 | 240 | 74.4 | 17.3 | 48.0 | 86.7 |
| Ogbera et al. [31] 2011 | 201 | 75.0 | 59.0 | 19.0 | 64.0 |
| Nsiah et al. [35] 2015 | 150 | 48.6 | 41.3 | 32.7 | 60.0 |
| Osei-Yeboah et al. [40] 2017 | 162 | 48.2 | 23.5 | 16.7 | 66.7 |
| Woyesa et al. [41] 2017 | 314 | 61.3 | 39.2 | 70.4 | 28.0 |
| Tadewos et al. [42] 2017 | 270 | 40.7 | 47.0 | 68.1 | 28.1 |
| Birarra et al. [44] 2018 | 256 | 53.5 | 67.2 | 68.8 | 43.4 |
| Biadgo et al. [43] 2018 | 159 | 43.4 | 32.7 | 62.3 | 55.4 |
| **Pooled prevalence [95% CI]** |  | **55.9 [47.6, 64.2]** | **43.3 [33.5, 53.2]** | **48.0 [35.2, 60.7]** | **54.8 [43.2, 66.4]** |

CI: Confidence Interval; HDL-c: High density lipoprotein cholesterol; TG: Triglyceride; NCEP-ATP III: National Cholesterol Education Program–Adult Treatment Panel III.

Whereas the overall pooled prevalence of MetS component by IDF criteria was as follows: central obesity 61.6% [95% CI: 47.9, 75.3], low HDL-c 49.9% [95% CI: 37.3, 62.6], hypertriglyceridemia 49.2% [95% CI: 34.1, 64.4] and hypertension 56.1% [95% CI: 46.7, 65.4] as summarized in table 5.

**Table 5:**
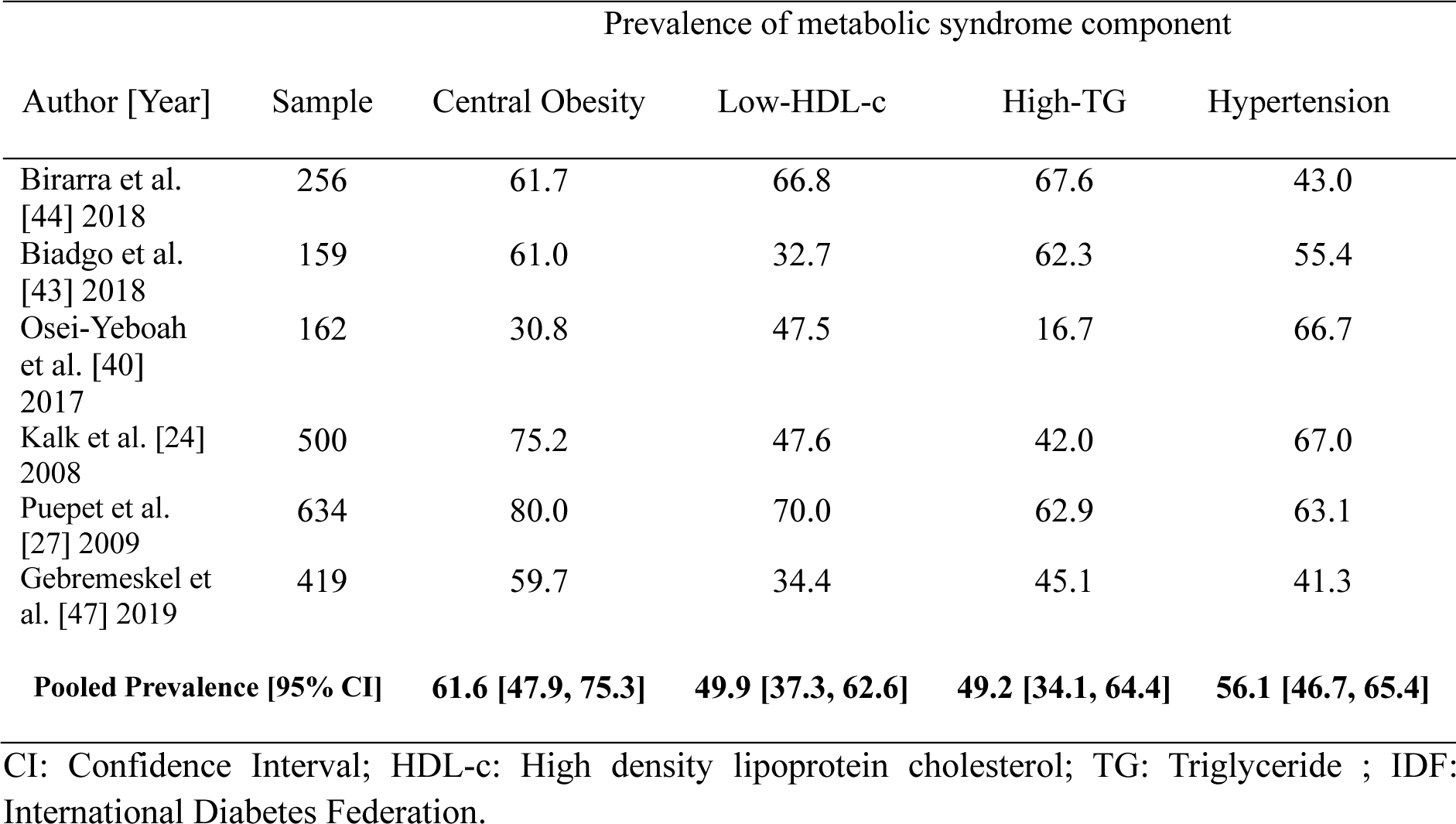
Pooled prevalence of metabolic syndrome component based on IDF criteria.

### Subgroup and sensitivity analysis

Subgroup analyses were conducted based on gender, country, sample size, and mean age. According to the NCEP-ATP III 2004, a total of 17 studies reported prevalence based on gender, revealing that the pooled prevalence of MetS among females in SSA was significantly higher compared to males (73.5% vs. 50.5%). Meanwhile, the results of subgroup analysis based on sample size showed the highest prevalence in studies with ≥250 subjects compared to those with <250 subjects (67.0% vs. 55.2%), as depicted in supplementary table 2. Furthermore, subgroup analysis based on IDF criteria, as shown in supplementary table 3, revealed a higher pooled prevalence among females (71.6%) compared to males (44.5%) among the 11 studies that reported prevalence based on gender. Among the 12 reports that specified participant mean age, the pooled prevalence was comparable across the two categories of mean age: <50 years and ≥50 years. Additionally, sensitivity analyses were conducted using the leave-one-out approach to evaluate the influence of individual studies on the overall estimate of MetS, based on the NCEP-ATP III 2004 and IDF criteria. The results indicated no substantial evidence for the influence of any single study on the overall pooled prevalence of MetS among individuals with T2DM in SSA (Figures 5 and 6). To further explore the observed heterogeneity in the study, we conducted a meta-regression to account for this. The analysis revealed that gender had a significant influence on the overall effect sizes in both NCEP-ATP III 2004 and IDF (p<0.0001, 0.0007 respectively) and studies with a sample size ≥ 250 for NCEP-ATP III 2004 there was a significant influence observed at p value 0.0106.

**Figure 5:**
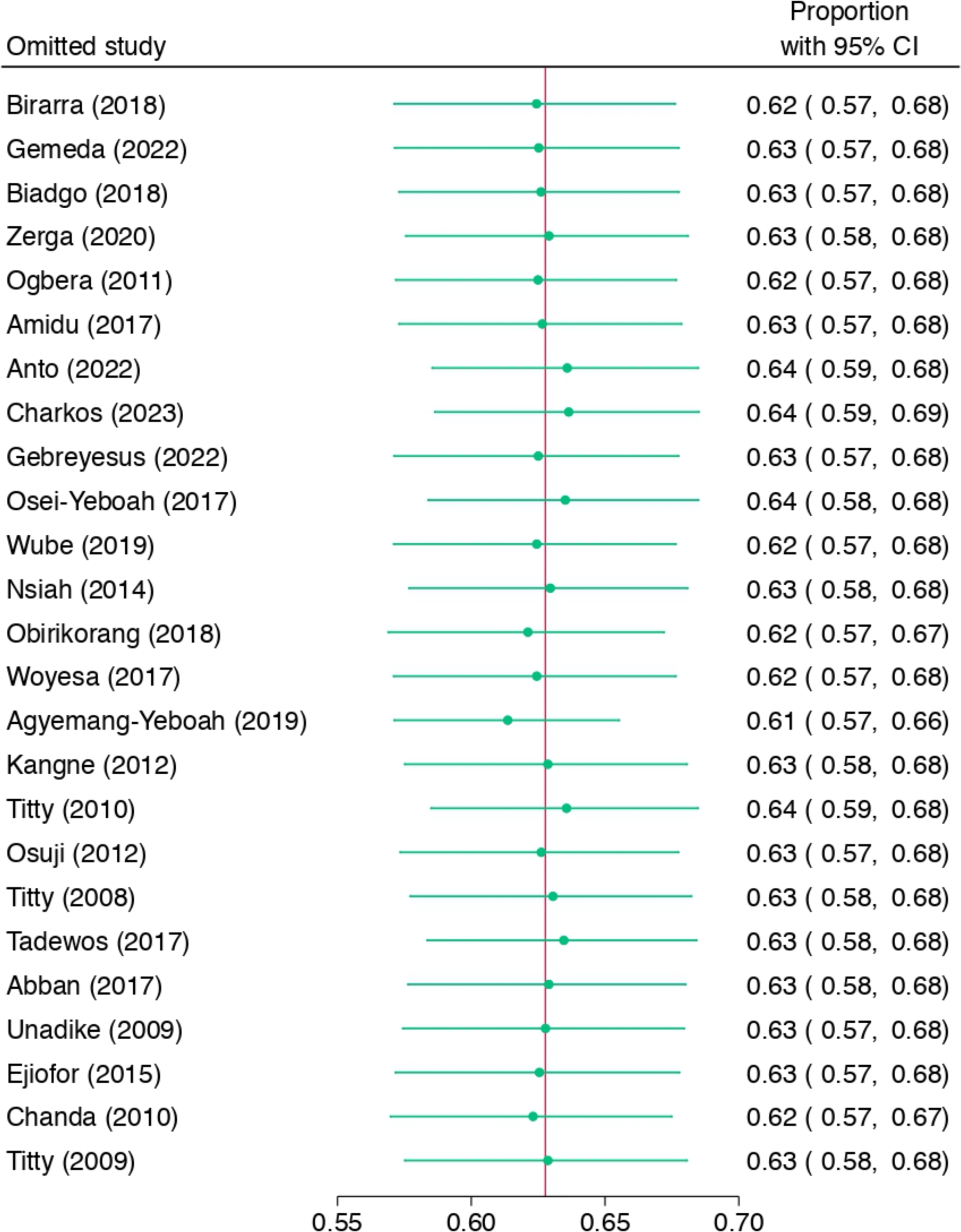
Sensitivity analysis based on NCEP-ATP III 2004 criteria.

**Figure 6:**
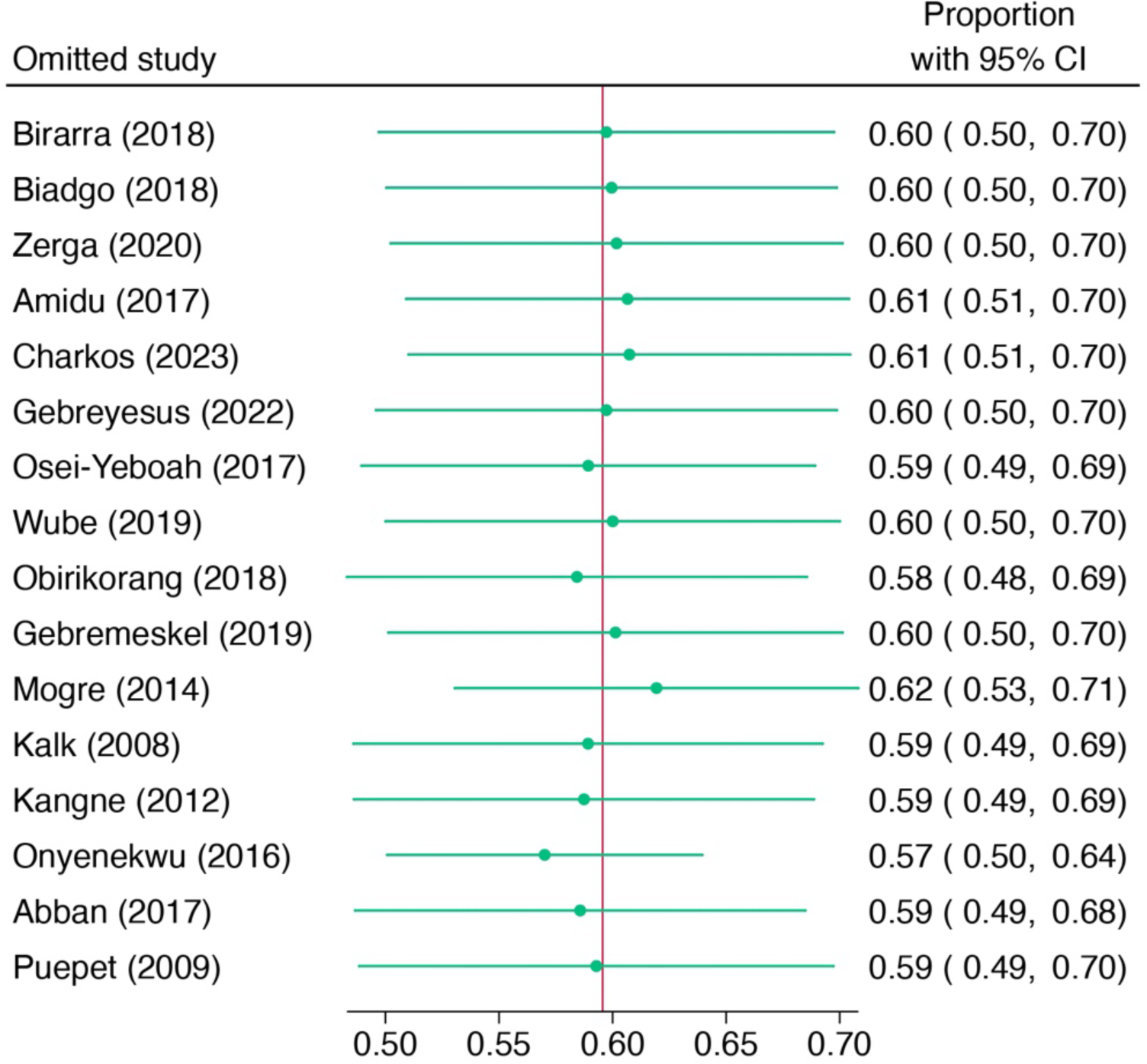
Sensitivity analysis based on IDF criteria.

### Publication bias

A funnel plot of the pooled prevalence of MetS and Begg’s statistical tests at a 5% significance level were used to assess the presence of potential publication bias among the included studies. The funnel plots were almost symmetrical for the NCEP-ATP III 2004 criteria and IDF criteria, as graphically represented in Figures 7 and 8, respectively. Furthermore, separate analyses of the linear regression test of funnel plot asymmetry based on NCEP-ATP III 2004 and IDF criteria resulted in statistically non-significant p-values of 0.7800 and 0.6686, respectively, indicating the absence of publication bias.

**Figure 7:**
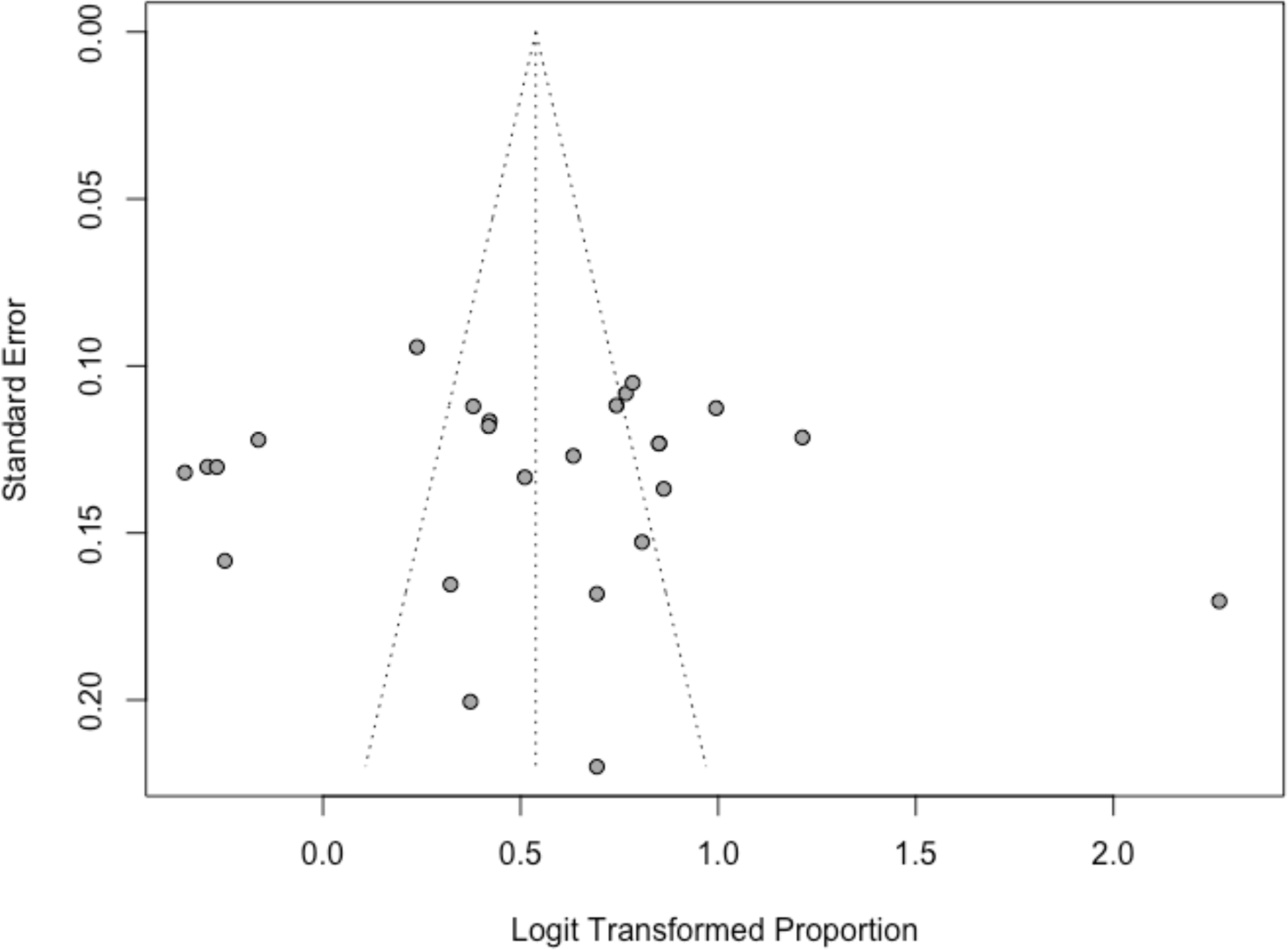
Funnel plot for the publication bias based on NCEP-ATP III 2004 criteria.

**Figure 8:**
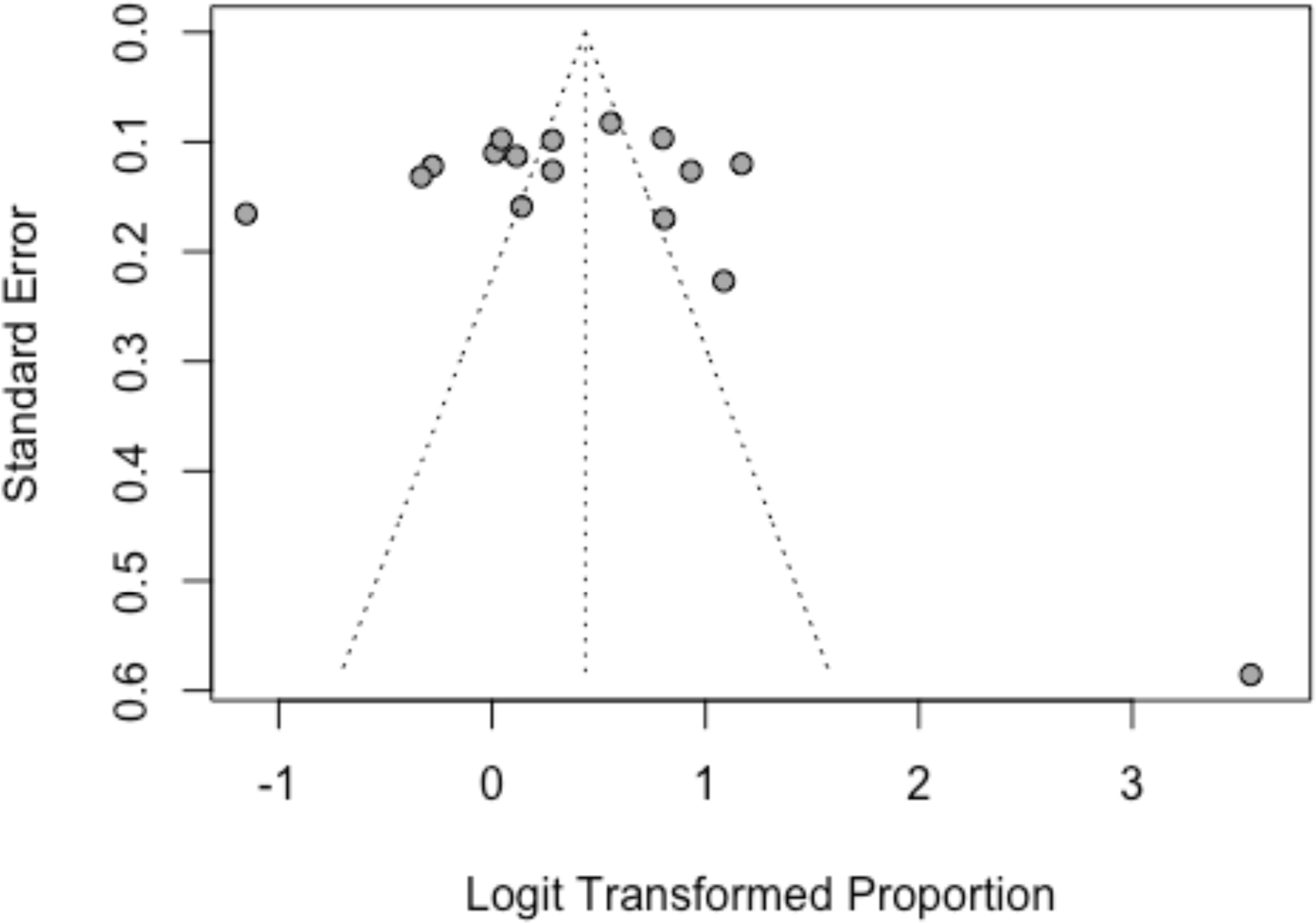
Funnel plot for the publication bias based on IDF criteria.

## Discussion

The association between T2DM and MetS has been thoroughly investigated. To our knowledge, this is the first systematic review and meta-analysis that evaluated the weighted pooled prevalence of MetS in individuals with T2DM in sub-Saharan Africa using specific diagnostic criteria for metabolic syndrome. The findings of this systematic review indicate that the weighted pooled prevalence of MetS was 63.1% (95% CI: 57.9 – 68.1) and 60.8% (95% CI: 50.7 – 70.0) using NCEP-ATP III 2004 and IDF criteria, respectively. The observed disparities in the prevalence of MetS when applying the NCEP-ATP III 2004 criteria versus the IDF criteria are noteworthy. The prevalence was slightly higher (63.1%) when the NCEP-ATP III 2004 criteria were used, compared to the IDF criteria (60.8%). These differences can be attributed to variations in the diagnostic components and thresholds employed by each set of criteria [57]. Similar findings regarding the variation in MetS prevalence based on diagnostic criteria have been reported in many studies conducted in different parts of the world [58,59]. Interestingly, when we compare our findings with those from other regions and study populations, we observe divergent outcomes. For instance, our findings are somewhat consistent with results reported in a systematic review among African T2DM patients (66.9%) [18] and Ethiopian T2DM patients (63.78%) [19]. However, the current weighted pooled prevalence of MetS using IDF criteria (60.8%) was higher than the prevalence estimated globally, which typically ranges between 20% and 25% when using similar diagnostic criteria [16].

Notably, subgroup analysis by gender revealed a considerably higher pooled prevalence of MetS in females, at 73.5% (95% CI: 67.4-79.5), compared to males at 50.5% (95% CI: 43.8-57.2) according to the NCEP-ATP III 2004. Similarly, a higher pooled prevalence was observed according to the IDF criteria among females, reaching 71.6% (95% CI: 60.2-82.9), compared to males at 44.5% (95% CI: 34.2-54.8). This finding aligns with reports from systematic reviews conducted among various populations, including SSA African [60], Ghanaian [61], Bangladesh [62], and mainland China [63]. The possible reason for the higher prevalence in females could be gender-specific increased MetS risk factors among women, such as menopause, contraceptive therapy use, elevated body weight, and increased waist circumference, in comparison to men [64]. Based on IDF criteria, among the included studies, the highest weighted pooled prevalence was observed in Nigeria at 80.2% (95% CI: 47.1-99.9), while Ethiopia had the lowest at 52.0% (95% CI: 48.3-55.8). This contrasts with a review by Shiferaw et al. [65]that identified the highest prevalence of MetS in Ethiopia. However, their study combined studies with varying diagnostic criteria, unlike our report, which might account for this variation. The differences in MetS prevalence between Nigeria and Ethiopia found on the current review stem from a blend of varying dietary patterns, lifestyle distinctions, disparities in healthcare infrastructure, and cultural influences.

Generally, our findings differ from those of many other studies around the world. In a systematic review conducted among healthy South Asians, a prevalence of MetS was reported as 26.1% (ATP III), 29.8% (IDF), and 32.5% (modified ATP III) [66]. Similarly, a quantitative synthesis of 111 studies conducted among the Indian adult general population reported a prevalence of 29% (NCEP ATP-III) and 34% (IDF) [67]. The observed discrepancies in the prevalence of MetS reported among different studies around the world are significant. These discrepancies might be due to differences in intrinsic study design, sample size, and characteristics of the study participants, such as comorbidities, geographical locations, urbanization, and lifestyle factors, including physical inactivity and unhealthy eating habits [68,69]. Moreover, the current review focused on Sub-Saharan African Type 2 Diabetes Mellitus individuals. T2DM appears to play a pivotal role in the pathogenesis and exacerbation of MetS, such that individuals with T2DM are more likely to have MetS, increasing their susceptibility to cardiovascular complications[11,70].

According to the data compiled in this review, the pooled prevalence of MetS components was as follows: central obesity at 55.9% and 61.6%; low HDL-c at 43.3% and 49.9%; hypertriglyceridemia at 48.0% and 49.2%; and hypertension at 54.8% and 56.1%, according to NCEP-ATP III 2004 and IDF criteria, respectively. Central obesity emerged as the most frequent metabolic syndrome component in this systematic review. Visceral adiposity has long been recognized as a central player in insulin resistance and is linked to a heightened risk of type 2 diabetes mellitus and cardiovascular diseases [71]. Moreover, high blood pressure and abnormal lipid profiles were also found to be prevalent in our review. Thus, our findings underscore the importance of a holistic approach to patient care, integrating strategies to mitigate MetS components alongside T2DM management to prevent adverse health effects such as CVD [72,73].

The strengths of the present study include its comprehensive database search using varying combinations of keywords and well-defined inclusion/exclusion criteria. However, we wish to acknowledge several limitations in the current study. Firstly, significant heterogeneity was observed across the included studies, and this heterogeneity persisted even after stratification for diagnostic criteria. Secondly, the diversity in sub-Saharan African populations, as SSA is home to various ethnic, cultural, and socio-economic groups, may exhibit different risk factors and disease profiles. Therefore, the generalizability of findings across this region may be limited, as the prevalence and associations of MetS in T2DM can vary among these subpopulations.

## Conclusion

Although limited in scope, the findings presented here underscore the alarming prevalence of MetS among individuals with T2DM in sub-Saharan Africa. This trend may be directly linked to the rapid economic development and urbanization occurring in the region. This swift industrialization can lead to significant changes in lifestyle patterns and overnutrition, resulting in overweight and obesity, emphasizing the urgent need for comprehensive, region-specific prevention and management strategies. Encouraging lifestyle modifications, including regular exercise and balanced diets, is essential. Moreover, it is crucial to develop routine obesity screening procedures. Implementing early interventions and robust public health initiatives are crucial in mitigate the risks associated with central obesity.

Sub-Saharan Africa faces unique health challenges, including limited healthcare resources and the dual burden of communicable and non-communicable diseases, which must be taken into account when developing effective interventions. Moving forward, it is imperative to prioritize research efforts that not only elucidate the underlying mechanisms of MetS and T2DM but also explore culturally sensitive and sustainable approaches for prevention and treatment. We hope that this systematic review will serve as a foundation for further studies, ultimately leading to more effective strategies and improved health outcomes for individuals in sub-Saharan Africa who are grappling with the challenges of metabolic syndrome and T2DM.

## Supporting information

Supplemental File 1: Tables for Database Search Strategy

Supplemental File 2: Tables for Subgroup Analysis

## Data Availability

All data produced in the present study are available upon reasonable request to the authors

## Acknowledgments

We would like to thank all authors of studies included in this systematic review and meta-analysis.

## Data Availability

The data used to support the findings of this study are available from the corresponding author upon request.

## Authors’ contributions

NM, HN and GJ developed the protocol and involved in the design, selection of study, data extraction, quality assessment, statistical analysis, results from interpretation, and developing the initial and final drafts of the manuscript. FS, CN, SG, AM, SH, KK and EM Involved in statistical analysis and revising subsequent drafts. All authors read and approved the final draft of the manuscript.

## Supplementary Materials

S1 Tables: Search Strategies used for final search of databases.

S2 Tables: Subgroup analysis results based on NCEP-ATP III 2004 and IDF criteria

