## Supplemental File 1: Tables for Database Search Strategy for "Exploring the Prevalence and Components of Metabolic Syndrome in Sub-Saharan African Type 2 Diabetes Mellitus Patients: A Systematic Review and Meta-Analysis"

**Search terms used for final search of databases, 18 August 2023**

**PubMed**

|  | **Search Strategy** | **Number of Hits** |
| --- | --- | --- |
| **#1** | "Metabolic Syndrome"[Mesh] OR "Metabolic Syndrome" [tw] OR "Metabolic Syndrome X"[tw] OR "Insulin Resistance Syndrome"[tw] OR "Cardiometabolic Syndrome" [tw] OR "Metabolic Disease*" [tw] OR "Metabolic Disorder*" [tw] OR "Abdominal Obesity" [tw] OR "Syndrome X" [tw] OR MetS [tw] | 145,463 |
| **#2** | "Diabetes Mellitus"[Mesh] OR “Type 2 diabetes mellitus” [tw] OR “Type 2 diabetes” [tw] OR T2DM[tw] OR “Diabetes Mellitus”[tw] OR “Non-insulin dependent diabetes”[tw] OR “Adult onset diabetes”[tw] OR Hyperglycemia[tw] OR “Insulin Resistance”[tw]. | 723,859 |
| **#3** | "Africa South of the Sahara"[Mesh] OR "Africa South of the Sahara" [tw] OR "Subsaharan Africa" [tw] OR "Sub-Saharan Africa" [tw] OR "sub Saharan Africa" [tw] OR "sub Saharan African" [tw] OR "Central Africa" [tw] OR "Central African" [tw] OR "West Africa" [tw] OR "West African" [tw] OR "Western Africa" [tw] OR "Western African" [tw] OR "East Africa" [tw] OR "East African" [tw] OR "Eastern Africa" [tw] OR "Eastern African" [tw] OR "South African" [tw] OR "Southern Africa" [tw] OR "Southern African" [tw] OR Angola[tw] OR Angolan[tw] OR Benin[tw] OR Botswana[tw] OR "Burkina Faso" [tw] OR Burundi[tw] OR "Cape Verde" [tw] OR Cameroon[tw] OR Cameroonian[tw] OR "Central African Republic" [tw] OR Chad[tw] OR Congo[tw] OR "Cote d'Ivoire" [tw] OR "Ivory Coast" [tw] OR "Democratic Republic of the Congo" [tw] OR Djibouti[tw] OR "Equatorial Guinea" [tw] OR Eritrea[tw] OR Eswatini[tw] OR Ethiopia[tw] OR Ethiopian[tw] OR Gabon[tw] OR Gabonese[tw] OR Gambia[tw] OR Gambian[tw] OR Ghana[tw] OR Ghanaian[tw] OR Guinea[tw] OR "Guinea-Bissau" [tw] OR Kenya[tw] OR Kenyan[tw] OR Lesotho[tw] OR Liberia[tw] OR Liberian[tw] OR Malawi[tw] OR Malawian[tw] OR Mali[tw] OR Mauritania[tw] OR Mauritanian[tw] OR Mozambique[tw] OR Namibia[tw] OR Namibian[tw] OR Niger[tw] OR Nigeria[tw] OR Nigerian[tw] OR Rwanda[tw] OR Rwandan[tw] OR "Sao Tome and Principe" [tw] OR Senegal[tw] OR Senegalese[tw] OR "Sierra Leone"[tw] OR Somalia[tw] OR "South Africa" [tw] OR "South Sudan" [tw] OR Sudan[tw] OR Sudanese[tw] OR Tanzania[tw] OR Tanzanian[tw] OR Togo[tw] OR Togolese[tw] OR Uganda[tw] OR Ugandan[tw] OR Zambia[tw] OR Zambian[tw] OR Zimbabwe[tw] OR Zimbabwean[tw] | 549,910 |
| **#4** | **#1 AND #2 AND #3** | **532** |

**Scopus Search Strategy**

|  | **Search Strategy** | **Number of Hits** |
| --- | --- | --- |
| **#1** | TITLE-ABS-KEY("Metabolic Syndrome" OR "Metabolic Syndrome X" OR "Insulin Resistance Syndrome" OR "Cardiometabolic Syndrome" OR "Metabolic Disease*" OR "Metabolic Disorder*" OR "Abdominal Obesity" OR "Syndrome X" OR MetS) | 232,626 |
| **#2** | TITLE-ABS-KEY ( "type 2 diabetes mellitus" OR "type 2 diabetes" OR "diabetes mellitus" OR "non-insulin dependent diabetes" OR "adult onset diabetes" OR hyperglycemia OR "insulin resistance" ) | 1,094,412 |
| **#3** | TITLE-ABS-KEY("Africa South of the Sahara" OR "Subsaharan Africa" OR "Sub-Saharan Africa" OR "sub Saharan Africa" OR "sub Saharan African" OR "Central Africa" OR "Central African" OR "West Africa" OR "West African" OR "Western Africa" OR "Western African" OR "East Africa" OR "East African" OR "Eastern Africa" OR "Eastern African" OR "South African" OR "Southern Africa" OR "Southern African" OR Angola OR Angolan OR Benin OR Botswana OR "Burkina Faso" OR Burundi OR "Cape Verde" OR Cameroon OR Cameroonian OR "Central African Republic" OR Chad OR Congo OR "Cote d'Ivoire" OR "Ivory Coast" OR "Democratic Republic of the Congo" OR Djibouti OR "Equatorial Guinea" OR Eritrea OR Eswatini OR Ethiopia OR Ethiopian OR Gabon OR Gabonese OR Gambia OR Gambian OR Ghana OR Ghanaian OR Guinea OR "Guinea-Bissau" OR Kenya OR Kenyan OR Lesotho OR Liberia OR Liberian OR Malawi OR Malawian OR Mali OR Mauritania OR Mauritanian OR Mozambique OR Namibia OR Namibian OR Niger OR Nigeria OR Nigerian OR Rwanda OR Rwandan OR "Sao Tome and Principe" OR Senegal OR Senegalese OR "Sierra Leone" OR Somalia OR "South Africa" OR "South Sudan" OR Sudan OR Sudanese OR Tanzania OR Tanzanian OR Togo OR Togolese OR Uganda OR Ugandan OR Zambia OR Zambian OR Zimbabwe OR Zimbabwean) | 1,159,988 |
| **#4** | **#1 AND #2 AND #3**  ( TITLE-ABS-KEY ( "Metabolic Syndrome" OR "Metabolic Syndrome X" OR "Insulin Resistance Syndrome" OR "Cardiometabolic Syndrome" OR "Metabolic Disease*" OR "Metabolic Disorder*" OR "Abdominal Obesity" OR "Syndrome X" OR mets ) ) AND ( TITLE-ABS-KEY ( hypertension OR "high blood pressure" ) ) AND ( TITLE-ABS-KEY ( "Africa South of the Sahara" OR "Subsaharan Africa" OR "Sub-Saharan Africa" OR "sub Saharan Africa" OR "sub Saharan African" OR "Central Africa" OR "Central African" OR "West Africa" OR "West African" OR "Western Africa" OR "Western African" OR "East Africa" OR "East African" OR "Eastern Africa" OR "Eastern African" OR "South African" OR "Southern Africa" OR "Southern African" OR angola OR angolan OR benin OR botswana OR "Burkina Faso" OR burundi OR "Cape Verde" OR cameroon OR cameroonian OR "Central African Republic" OR chad OR congo OR "Cote d'Ivoire" OR "Ivory Coast" OR "Democratic Republic of the Congo" OR djibouti OR "Equatorial Guinea" OR eritrea OR eswatini OR ethiopia OR ethiopian OR gabon OR gabonese OR gambia OR gambian OR ghana OR ghanaian OR guinea OR "Guinea-Bissau" OR kenya OR kenyan OR lesotho OR liberia OR liberian OR malawi OR malawian OR mali OR mauritania OR mauritanian OR mozambique OR namibia OR namibian OR niger OR nigeria OR nigerian OR rwanda OR rwandan OR "Sao Tome and Principe" OR senegal OR senegalese OR "Sierra Leone" OR somalia OR "South Africa" OR "South Sudan" OR sudan OR sudanese OR tanzania OR tanzanian OR togo OR togolese OR uganda OR ugandan OR zambia OR zambian OR zimbabwe OR zimbabwean ) ) AND ( LIMIT-TO ( LANGUAGE , "English" ) ) | **850** |

**EMBASE Search Strategy With (title, Abstract and Keywords)**

|  | **Search Strategy** | **Number of Hits** |
| --- | --- | --- |
| **#1** | 'metabolic syndrome x':ti,ab,kw OR 'cardiometabolic syndrome':ti,ab,kw OR 'cardiometabolic disease':ti,ab,kw OR 'abdominal obesity':ti,ab,kw OR 'metabolic disorder':ti,ab,kw | 30,661 |
| **#2** | ('non insulin dependent diabetes mellitus' OR 'diabetes mellitus' OR hyperglycemia OR 'insulin resistance'):ti,ab,kw | 553,718 |
| **#3** | (('africa south of the sahara':ti,ab,kw OR 'sub-saharan african':ti,ab,kw OR 'west africa':ti,ab,kw OR 'west african':ti,ab,kw OR 'central africa':ti,ab,kw OR 'central african':ti,ab,kw OR 'east african':ti,ab,kw OR 'south african':ti,ab,kw OR angola:ti,ab,kw OR angolan:ti,ab,kw OR benin:ti,ab,kw OR beninese:ti,ab,kw OR burundi:ti,ab,kw OR botswana:ti,ab,kw OR 'burkina faso':ti,ab,kw OR burkinabe:ti,ab,kw OR 'cape verde':ti,ab,kw OR cameroon:ti,ab,kw OR cameroonian:ti,ab,kw OR 'central african republic':ti,ab,kw OR chad:ti,ab,kw OR congo:ti,ab,kw OR 'cote d`ivoire':ti,ab,kw OR 'democratic republic congo':ti,ab,kw OR djibouti:ti,ab,kw OR 'equatorial guinea':ti,ab,kw OR eritrea:ti,ab,kw OR eritrean:ti,ab,kw OR eswatini:ti,ab,kw OR ethiopia:ti,ab,kw OR ethiopian:ti,ab,kw OR gabon:ti,ab,kw OR gabonese:ti,ab,kw OR gambia:ti,ab,kw OR gambian:ti,ab,kw OR ghana:ti,ab,kw OR ghanaian:ti,ab,kw OR guinea:ti,ab,kw OR 'guinea bissau':ti,ab,kw OR kenya:ti,ab,kw OR kenyan:ti,ab,kw OR lesotho:ti,ab,kw OR liberia:ti,ab,kw OR liberian:ti,ab,kw OR malawi:ti,ab,kw OR malawian:ti,ab,kw OR mali:ti,ab,kw OR mauritania:ti,ab,kw OR mauritanian:ti,ab,kw OR mozambique:ti,ab,kw OR mozambican:ti,ab,kw OR namibia:ti,ab,kw OR namibian:ti,ab,kw OR niger:ti,ab,kw OR nigeria:ti,ab,kw OR nigerian:ti,ab,kw OR rwanda:ti,ab,kw OR rwandan:ti,ab,kw OR 'sao tome':ti,ab,kw) AND principe:ti,ab,kw OR senegal:ti,ab,kw OR senegalese:ti,ab,kw OR 'sierra leone':ti,ab,kw OR somalia:ti,ab,kw OR somali:ti,ab,kw) AND citizen:ti,ab,kw OR 'south africa':ti,ab,kw OR 'south sudan':ti,ab,kw OR sudan:ti,ab,kw OR sudanese:ti,ab,kw OR tanzania:ti,ab,kw OR tanzanian:ti,ab,kw OR togo:ti,ab,kw OR togolese:ti,ab,kw OR uganda:ti,ab,kw OR ugandan:ti,ab,kw OR zambia:ti,ab,kw OR zambian:ti,ab,kw OR zimbabwe:ti,ab,kw OR zimbabwean:ti,ab,kw | 122,726 |
| **#4** | **#1 AND #2 AND #3** | **26** |
