## Supplemental File 2: Tables for Subgroup Analysis for "Exploring the Prevalence and Components of Metabolic Syndrome in Sub-Saharan African Type 2 Diabetes Mellitus Patients: A Systematic Review and Meta-Analysis"

Table 6: Subgroup meta-analysis of metabolic syndrome prevalence among T2DM patients in SSA based on NCEP-ATP III 2004.

| **Category** | **Subgroup** | **Number of studies** | **Sample size** | **Prevalence (95% CI)** | **Heterogeneity** | | |
| --- | --- | --- | --- | --- | --- | --- | --- |
|  |  |  |  |  | **Q Statistic** | ***I^2^*** | **p-value** |
| Country | Cameroon | 1 | 308 | 60.4% | - | - | - |
|  | Ethiopia | 9 | 2695 | 62.4 (55.2-69.6) | 112.4 | 93.9% | <0.001 |
|  | Ghana | 10 | 2715 | 59.8 (50.2-69.5) | 470.0 | 97.0% | <0.001 |
|  | Nigeria | 4 | 900 | 66.6 (63.6-69.7) | 2.6 | 0% | 0.464 |
|  | Zambia | 1 | 400 | 73.0% | - | - | - |
| Gender | Male | 17 | 2182 | 50.5 (43.8-57.2) | 182.6 | 90.8% | <0.001 |
|  | Female | 17 | 2833 | 73.5 (67.4-79.5) | 314.6 | 94.3% | <0.001 |
| Sample Size | ≥250 | 15 | 5192 | 67.0 (61.8-72.2) | 357.6 | 94.5% | <0.001 |
|  | <250 | 10 | 1826 | 55.2 (48.1-62.3) | 95.4 | 89.9% | <0.001 |
| Mean Age | < 50 Year | 5 | 1297 | 59.3 (47.5-71.0) | 82.0 | 95.1% | <0.001 |
|  | ≥50 Years | 14 | 3897 | 65.3 (59.3-71.2) | 341.4 | 94.2% | <0.001 |

Table 7: Subgroup meta-analysis of metabolic syndrome prevalence among T2DM patients in SSA based on IDF criteria

| **Category** | **Subgroup** | **Number of studies** | **Sample size** | **Prevalence (95% CI)** | **Heterogeneity** | | |
| --- | --- | --- | --- | --- | --- | --- | --- |
|  |  |  |  |  | **Q Statistic** | ***I^2^*** | **p-value** |
| Country | Cameroon | 1 | 308 | 71.7 | - | - | - |
|  | Ethiopia | 7 | 2136 | 52.0 (48.3-55.8) | 17.8 | 67.5% | 0.007 |
|  | Ghana | 5 | 1123 | 57.4 (37.2-77.6) | 247.4 | 98.2% | <0.001 |
|  | Nigeria | 2 | 742 | 80.2 (47.1-99.9) | 184.1 | 99.5% | <0.001 |
|  | South Africa | 1 | 500 | 69.0 | - | - | - |
| Gender | Male | 11 | 1690 | 44.5 (34.2-54.8) | 233.0 | 95.2% | <0.001 |
|  | Female | 11 | 2067 | 71.6 (60.2-82.9) | 390.8 | 97.7% | <0.001 |
| Sample Size | ≥250 | 10 | 3840 | 59.3 (52.7-65.9) | 164.3 | 94.7% | <0.001 |
|  | <250 | 6 | 969 | 60.1 (39.2-81.0) | 618.6 | 98.7% | <0.001 |
| Mean Age | < 50 Years | 3 | 973 | 58.7 (48.1-69.4) | 26.6 | 90.8% | <0.001 |
|  | ≥50 Years | 9 | 2905 | 58.9 (47.7-70.2) | 288.2 | 97.6% | <0.001 |
